# Deep phylogenetic-based clustering analysis uncovers new and shared mutations in SARS-CoV-2 variants as a result of directional and convergent evolution

**DOI:** 10.1101/2021.10.14.21264474

**Authors:** Danilo Rosa Nunes, Carla Torres Braconi, Louisa F. Ludwig-Begall, Clarice Weis Arns, Ricardo Durães-Carvalho

**Affiliations:** Department of Microbiology, Immunology and Parasitology, Paulista School of Medicine, Federal University of São Paulo, São Paulo-SP, Brazil; Veterinary Virology and Animal Viral Diseases, Department of Infectious and Parasitic Diseases, FARAH Research Centre, Faculty of Veterinary Medicine, University of Liège, Belgium; Laboratory of Virology, University of Campinas, Campinas-SP, Brazil

## Abstract

Nearly two decades after the last epidemic caused by a severe acute respiratory syndrome coronavirus (SARS-CoV), newly emerged SARS-CoV-2 quickly spread in 2020 and precipitated an ongoing global public health crisis. Both the continuous accumulation of point mutations, owed to the naturally imposed genomic plasticity of SARS-CoV-2 evolutionary processes, as well as viral spread over time, allow this RNA virus to gain new genetic identities, spawn novel variants and enhance its potential for immune evasion. Here, through an in-depth phylogenetic clustering analysis of upwards of 200,000 whole-genome sequences, we reveal the presence of not previously reported and hitherto unidentified mutations and recombination breakpoints in Variants of Concern (VOC) and Variants of Interest (VOI) from Brazil, India (Beta, Eta and Kappa) and the USA (Beta, Eta and Lambda). Additionally, we identify sites with shared mutations under directional evolution in the SARS-CoV-2 Spike-encoding protein of VOC and VOI, tracing a heretofore-undescribed correlation with viral spread in South America, India and the USA. Our evidence-based analysis provides well-supported evidence of similar pathways of evolution for such mutations in all SARS-CoV-2 variants and sub-lineages. This raises two pivotal points: the co-circulation of variants and sub-lineages in close evolutionary environments, which sheds light onto their trajectories into convergent and directional evolution (i), and a linear perspective into the prospective vaccine efficacy against different SARS-CoV-2 strains (ii).

**Author summary:** In this study, through analysis of very robust and comprehensive datasets, we identify a plethora of mutations in the SARS-CoV-2 Spike cell surface protein of several variants of concern and multiple variants of interest. We trace an association of such mutations with viral spread in different countries. We further infer the presence of new SARS-CoV-2 sublineages and show that the vast majority of mutations identified in the SARS-CoV-2 Spike protein are under convergent evolution. If we consider every color of a Rubik’s cube’s face to represent a different mutation of a particular variant, evolutionary convergence can be achieved only when all composite pieces of a single face are of the same color and every face has one unique color. Overall, this raises two important points: we provide insight into the presence of SARS-CoV-2 variants and sub-lineages circulating in very close evolutionary environments and our analyses can serve to facilitate an outlook into the prospective vaccine efficacy against different SARS-CoV-2 strains.

## Introduction

In the last two decades, human health has been threatened by the emergence of three important zoonotic and pathogenic betacoronaviruses, namely the severe acute respiratory syndrome coronavirus (SARS-CoV) (Guan et al., 2003), the Middle East respiratory syndrome coronavirus (MERS-CoV) (Zaki et al., 2012) and, most recently, the causative agent of the Coronavirus Disease 2019 (COVID-19) pandemic, SARS-CoV-2 (da Costa, Moreli, and Saivish 2020). Likely originated from bats, pandemic SARS-CoV-2, like other endemic human alpha- (NL63 and 229E) and beta- (OC43 and HKU1) CoVs known for causing upper respiratory tract infections, overcame the interspecies barrier as a result of spillover and/or recombination events, and gained a pervasive ability to rapidly infect and spread around the globe (Corman et al., 2018; Boni et al., 2020; V’kovski et al., 2021).

The COVID-19 pandemic precipitated an intense genomic surveillance via data depositories and sequencing platforms and led to an unprecedented accumulation of public genomic data concerning a human pathogenic virus (Boni et al., 2020; Munnink et al., 2021). The sheer amount of available sequencing data has the potential to facilitate higher-precision micro-evolutionary analyses mapping escape and point mutations in presumed positively selected sites and residues putatively associated to an increased virus fitness and pathogenesis and allows inferences concerning the dynamics of SARS-CoV-2 spread (Kosakovsky Pond et al., 2008; Alteri et al., 2021).

Although the analysis of micro-evolutionary mechanisms is of paramount importance and may provide powerful information to promote the prediction of vaccination perspectives and the tracing of SARS-CoV-2 epidemiological chains, there is as yet a lack of data-based investigations examining the presence of eventual shared mutations and their evolutionary characteristics in classified SARS-CoV-2 Variants of Concern (VOC) and Variants of Interest (VOI) (CDC 2021a; Peacock et al., 2021).

Given the importance of monitoring mutations to track the emergence of novel variants, here we investigate the influence of directional selection and the dynamics of SARS-CoV-2 genomic plasticity in VOC and VOI by clustering partition high-scale phylogenetic and directional evolution (DEPS) approaches. Additionally, we show the presence of several mutations common for both VOI/VOC and convergently emerged sub-lineages, and provide a perspective of possible effects on the vaccination efficacy and the ongoing COVID-19 pandemic.

## Methods

### Sequence data and filtering strategy

High-coverage and complete HCoV-229E and HCoV-NL63 (alpha-CoVs), HCoV-OC43, HCoV-HKU1, MERS-CoV, SARS-CoV and SARS-CoV-2 VOC and VOI (beta-CoVs) genome sequences (≥ 29,000 bp), sampled from humans, were retrieved from the Global Initiative on Sharing Avian Influenza Data-EpiCoV (GISAID-EpiCoV) and GenBank databases at different times: February 12^th^ (MERS-CoV, SARS-CoV and SARS-CoV-2), July 12^th^ (HCoV-229E, HCoV-NL63, HCoV-OC43, HCoV-HKU1 and SARS-CoV-2) and August 26^th^ 2021 (SARS-CoV-2), totalling 238,990 sequences. With regards to SARS-CoV-2, we particularly focused on strains of countries from South America, China, India, and the United States of America (USA). At the time of analysis, India, the USA, and Brazil had reported the largest numbers of cumulative confirmed COVID-19 cases and deaths. This approach was used to compare putative mutual sites and residue changes under directional evolution over time.

Subsequently, sequences were filtered via Sequence Cleaner, a biopython-based program, utilising the following script: sequence_cleaner -q INPUT_DIRECTORY -o OUTPUT_DIRECTORY -ml 29,000 (MINIMUM_LENGTH) - mn 0 (PERCENTAGE_N) --remove_ambiguous. The outcome was a set of unambiguous sequences equal to and greater than 29,000 pb with zero percent of unknown nucleotides. Next, the datasets were aligned by adding coding-sequences related to references for HCoV-229E (NC_002645.1), HCoV-NL63 (NC_005831.2), HCoV-OC43 (NC_006213.1), HCoV-HKU1 (NC_006577.2), MERS-CoV (NC_038294.1), SARS-CoV (NC_004718.3), and SARS-CoV-2 (NC_045512.2), using default settings, with the rapid calculation of full-length multiple sequence alignment of closely-related viral genomes (MAFFT v.7 web-version program; https://mafft.cbrc.jp/alignment/software/closelyrelatedviralgenomes.html) and were edited by the UGENE v.38.1 (Okonechnikov et al., 2012).

### Clustering and sub-clustering analysis

A methodological approach to extract large-scale phylogenetic partitions was applied to identify transmission cluster chains on the largest Maximum Likelihood (ML) phylogenetic trees of the SARS-CoV-2 variants on the basis of a depth-first search algorithm which unifies evaluation of node reliability, tree topology and patristic distance (Prosperi et al., 2011). The ML tree was implemented in FastTree v.2.1.7 by using the standard implementation General Time Reversible (GTR) plus CAT with 20 gamma distribution parameters and a mix of Nearest-Neighbor Interchanges (NNI) and Sub-Tree-Prune-Regraft (SPR) (Price, Dehal, and Arkin 2010). Thereafter, in view of identifying SARS-CoV-2 cluster transmission events, we first selected sequences (one per cluster) from nodes/sub-trees with ≥ 2 distinct individuals and ≥ 90% reliability of statistical support (Shimodaira-Hasegawa test), where initially the patristic distance was adjusted to find a representative number of clusters (n= 100) from each large reconstructed ML tree. In addition to this strategy, a second approach included sub-clustering analysis as an indirect way to infer and investigate the possibility of co-circulating sub-lineages. For this, we selected sequences (two per cluster) with ≥ 95% node reliability of statistical support from a threshold of 0.05, thus corresponding to the 5^th^ percentile when considering the whole-tree patristic distance distribution.

### Recombination and directional evolution analyses

Before proceeding to directional evolution analysis, all datasets were submitted to the Genetic Algorithm for Recombination Detection (GARD), a likelihood-based tool to pinpoint recombination breakpoints (Kosakovsky Pond et al., 2006). To double check the outcome of the first of the two strategies described above, an additional test was conducted using the Pairwise Homoplasy Index (PHI; default settings) (Huson and Bryant 2005). Evidence-based analysis through phylogenetic maximum-likelihood was then performed implementing the Datamonkey web-server and the program Hyphy v.2.5 to track directional selection in amino acid sequences (DEPS) (Kosakovsky Pond et al., 2020). The DEPS method identifies both the residue and sites evolving toward it with great accuracy and detects frequency-dependent selection-scenarios as well as selective sweeps and convergent evolution that can confound most existing tests (Kosakovsky Pond et al., 2008). Further, the DEPS method has shown better performance than (traditional) substitution rate-based analyses (dN/dS) in detecting transient and frequency-dependent selection and directionally evolving sites and residues. For the most part, a Beta-Gamma site-to-site rate variation was used to conduct the analysis. The best-fit protein substitution model was chosen according to the corrected Akaike Information Criterion (cAIC). Only target sites and residues with Empirical Bayes Factors for evidence in favour of a directional selection model equal to or greater than 100 were considered for further exploration. Certain randomly chosen datasets were run multiple times (more than eight) to confirm obtained results.

### Statistical analysis

Data pertaining to SARS-CoV and MERS-CoV-related cases and deaths were extracted from the National Health Service (NHS, UK) (https://www.nhs.uk/conditions/sars/) and European Centre for Disease Prevention and Control (ECDC) (https://www.ecdc.europa.eu/en/publications-data/distribution-confirmed-cases-mers-cov-place-infection-and-month-onset-1), respectively. Information concerning SARS-CoV-2 was collected from World Health Organization (WHO) (https://covid19.who.int/). Population demographic data were retrieved from the Our World in Data website (https://ourworldindata.org/grapher/world-population-by-world-regions-post-1820?tab=table&country=Oceania~~North+America~Europe~Africa~Asia).

Statistical analyses were performed using one-way analysis of variance (ANOVA) and nonparametric methods followed by *post hoc* Kruskal-Wallis and Friedman (both with Dunn’s Multiple Comparison), and Bartlett’s tests (Tukey’s, Newman-Keuls and Bonferroni’s multiple comparisons). Additionally, Mann Whitney and Wilcoxon matched-pairs signed-rank (T test) and Pearson/Spearman (Correlation), all one-tailed methods with 99% confidence interval (CI), were run. *P-values* equal to or less than 0.005 (*p* ≤ 0.005, SARS-CoV-2 from South America: DEPS [sites and residues] vs infections) and 0.05 (*p* ≤ 0.05, SARS-CoV-2 from Brazil, China, India and the USA: DEPS [residues] vs circulating variants and infections) were considered as statistically significant. Data analyses were carried out using GraphPad Prism v. 5.01 (GraphPad Software, San Diego, California, USA). Figures and data visualization were performed using the ggplot2 v.3.3.5 package in the R (RStudio v.1.4.1717) language environment. Final graphics were edited with the open-source software drawing tool Inkscape v.1.0.2.

## Results and discussion

Recombination is known to be a crucial evolutionary process for many RNA viruses (Lai 1992; Lemey, Salemi, and Vandamme 2009; Su et al., 2016); the process is frequently observed in the *Coronaviridae* family where recombination is likely facilitated by discontinuous transcription involving jumps of the replication-transcription complex during minus strand RNA synthesis. However, the consequences of recombination events occurring in the context of the current SARS-CoV-2 evolutionary landscape are still speculative (Li et al., 2020; Singh and Yi 2021; Pollett et al., 2021). Here we address this knowledge gap, revealing the presence of recombination and shared mutations in the SARS-CoV-2 Spike-encoding protein, demonstrating them to be under directional and convergent evolution amongst SARS-CoV-2 VOC/VOI and sub-lineages, and tracing an interconnection with viral spread. First, endemic and epidemic human coronaviruses (HCoVs) were compared to identify similar evolutionary patterns that could help clarify the evolution of SARS-CoV-2. An initial recombination breakpoint analysis showed that four of six HcoVs analyzed presented such signals (Fig. 1A). Endemic viruses OC43, NL63 and HKU1 also showed a similar pattern of residue accumulation and directional evolution, despite these viruses being subject to differing selective pressures (Forni et al., 2021).

**Figure 1.**
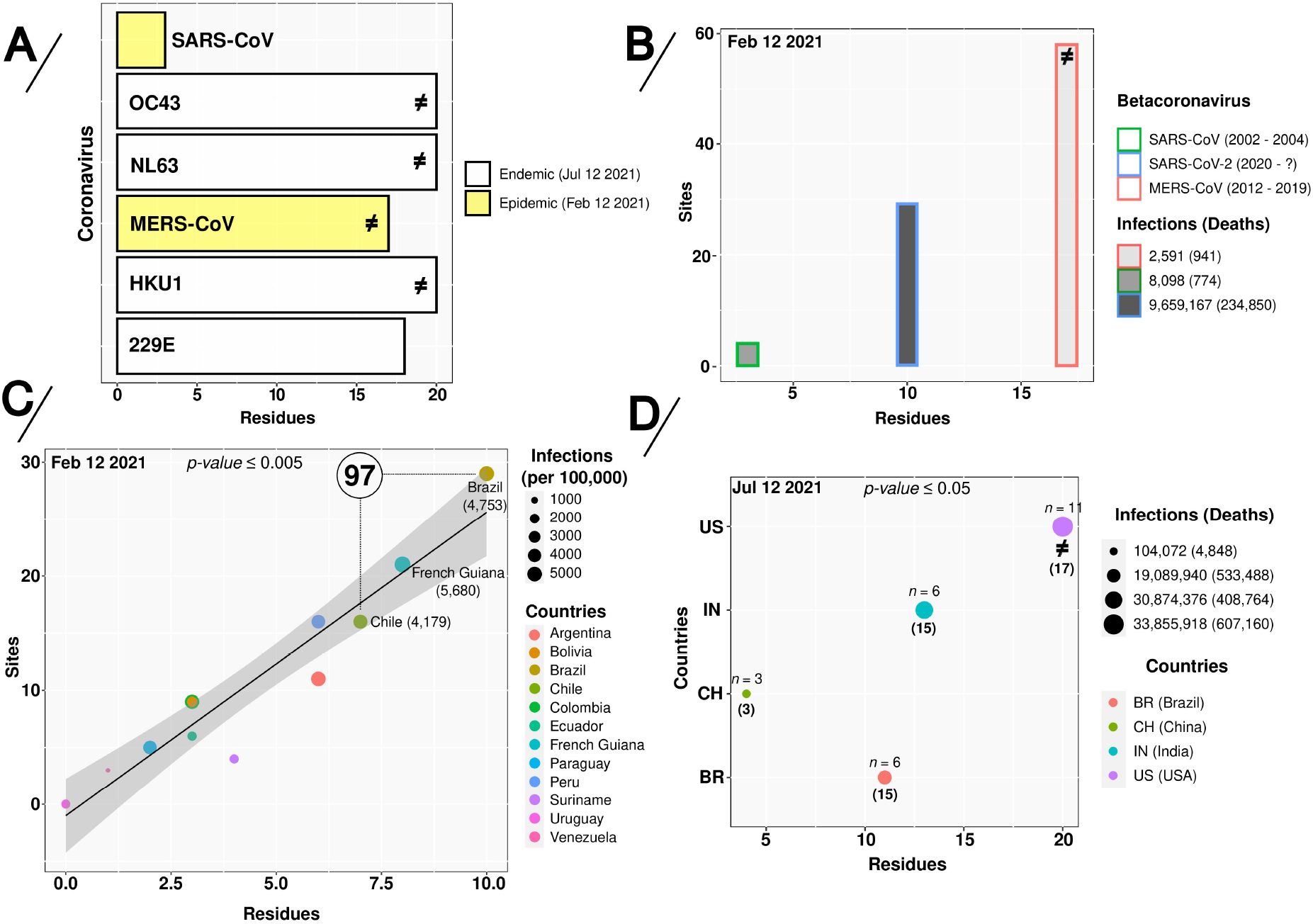
Directionally-evolving sites and residues in alpha- (229E and NL63) and beta- (OC43, HKU1, SARS-CoV, MERS-CoV and SARS-CoV-2) coronavirus sequences. (**A**) Directed-evolving residues in six different endemic and epidemic human coronaviruses (HCoVs), (**B**) Sites and residues vs infections in epidemic and pandemic coronaviruses (CoVs). (**C**) Linear-regression curve and co-relationship on the absolute amount of sites and/or residues under directional-positive selection given SARS-CoV-2 infections per 100,000 people from South America countries and (**D**) the total number of SARS-CoV-2 infections in Brazil, China, India and the USA. In panels A and B, the symbol ≠ represents the presence of recombination breakpoints signals. In panel C, the number inside the circle represents the amount of clusters found in Brazil and Chile. In panel D, *n* represents the amount of SARS-CoV-2 variants and the numbers in parentheses indicate sites under directional and convergent evolution in the Spike-encoding protein. Colors and symbols used in the panels are defined in the legend to the right of the figure.

A subsequent comparison of SARS-CoV-2 to the other two pathogenic HCoVs (SARS-CoV and MERS-CoV), highlighted differences in the number of directionally-evolving sites and residues (Fig. 1B). These patterns, putatively reflecting the initial evolutionary paths of the individual viruses, may suggest that SARS-CoV was initially under lower positive evolutionary pressure than MERS-CoV and SARS-CoV-2. In turn, deletions and mutations acquired by SARS-CoV have been shown to have had an impact on adaptation to human-to-human transmission, modifying both the capacity for viral proliferation and profiles of pathogenesis (Muth et al.,2018; Pereira 2020; Pereira 2021).

The evolution of SARS-CoV-2 was initially marked by genetic drift in a typical process of neutral evolution (Dearlove et al., 2020; MacLean et al., 2021); the virus reached a large number of new and susceptible hosts and, although some mutations appeared along the genome, there was no significant shift (Martin et al., 2021). However, as SARS-CoV-2 spread (Yadav et al., 2020; Hodcroft et al., 2021), fitness changes resulting from mutations in the viral genome as well as the emergence of new variants were increasingly reported (Velazquez-Salinas et al., 2020; Zhang et al., 2020; Plante et al., 2021).

The first epidemic wave of SARS-CoV-2 severely affected most countries in South America as a probable result of multiple viral introductions (Candido 2020); rapid increases of case numbers were especially reported in Brazil, the biggest and most populous country in Latin America (Paiva et al., 2020; Stefanelli et al., 2020). The uncontrolled viral spread created a favorable scenario for the emergence of new variants (Voloch et al., 2021; Faria et al., 2021; Resende et al., 2021; Sabino et al., 2021). To identify the impact of directional-positive selection sites at the rate of infections under these particular conditions, we traced the evolutionary scenario of SARS-CoV-2 in South America (via analysis of a significant and representative amount of genome sequences).

Remarkably, our data showed that an increase of DEPS was correlated with viral spread dynamics, with Brazil exhibiting a lower proportion of COVID-19 cases when compared to French Guiana and the same amount of SARS-CoV-2 clusters inferred in Chile (n= 97) (Fig. 1C), probably due to a higher diversity of circulating viruses. Our results also highlighted a series of mutations; while certain mutations have previously been described, but have hitherto remained unidentified in SARS-CoV-2 VOC and VOI, multiple further mutations are identified for the first time in this study (Table 1).

**Table 1:**
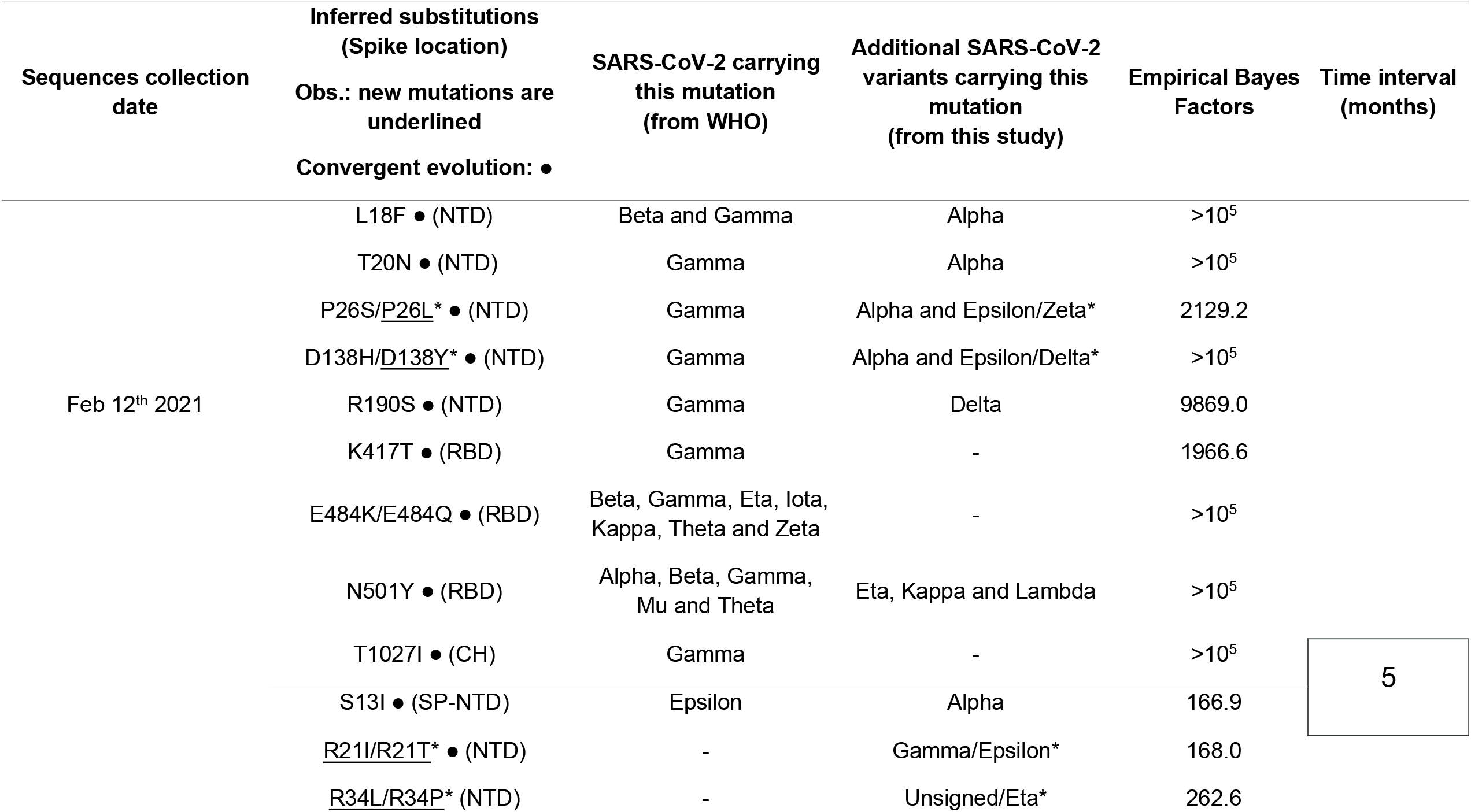
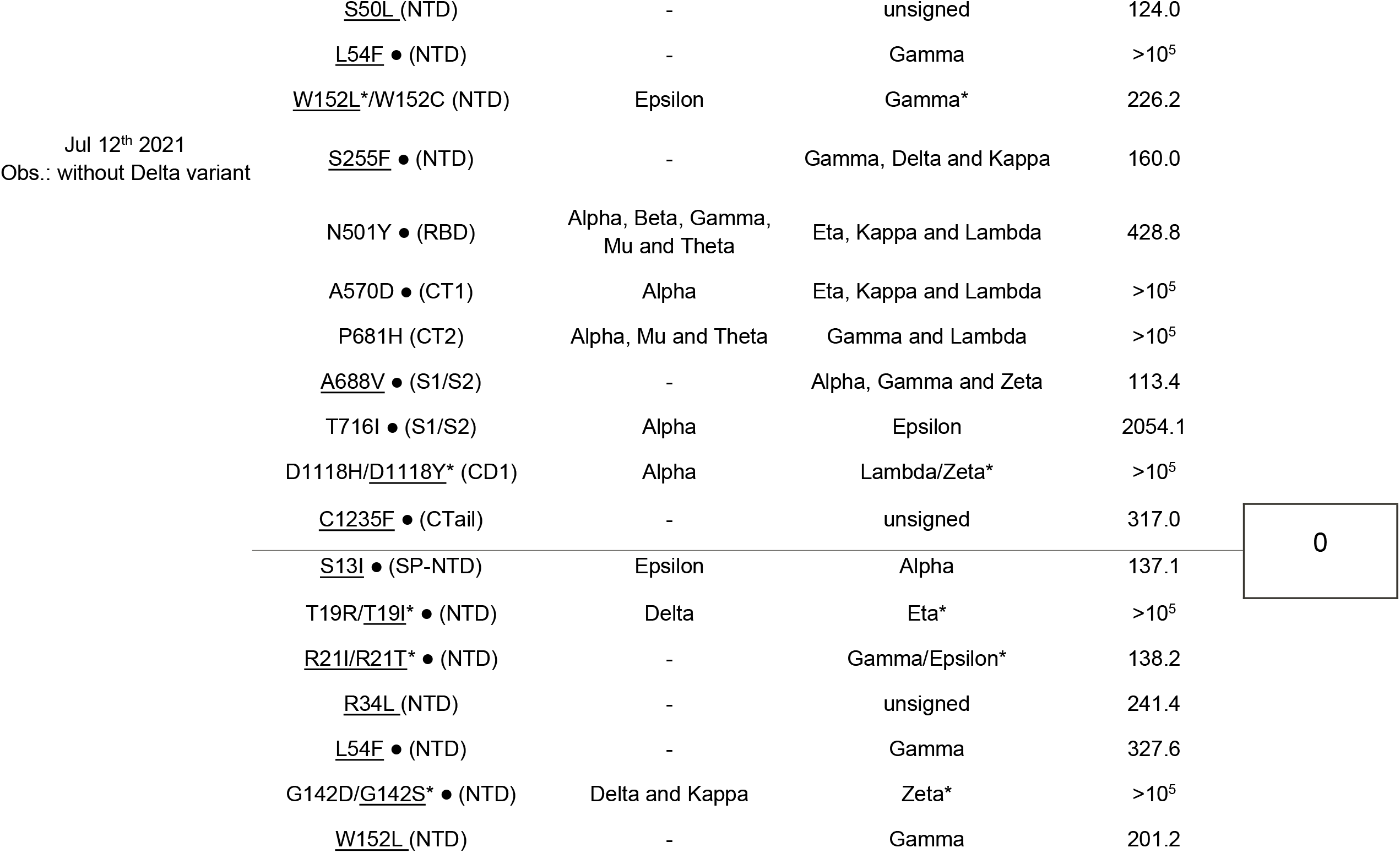
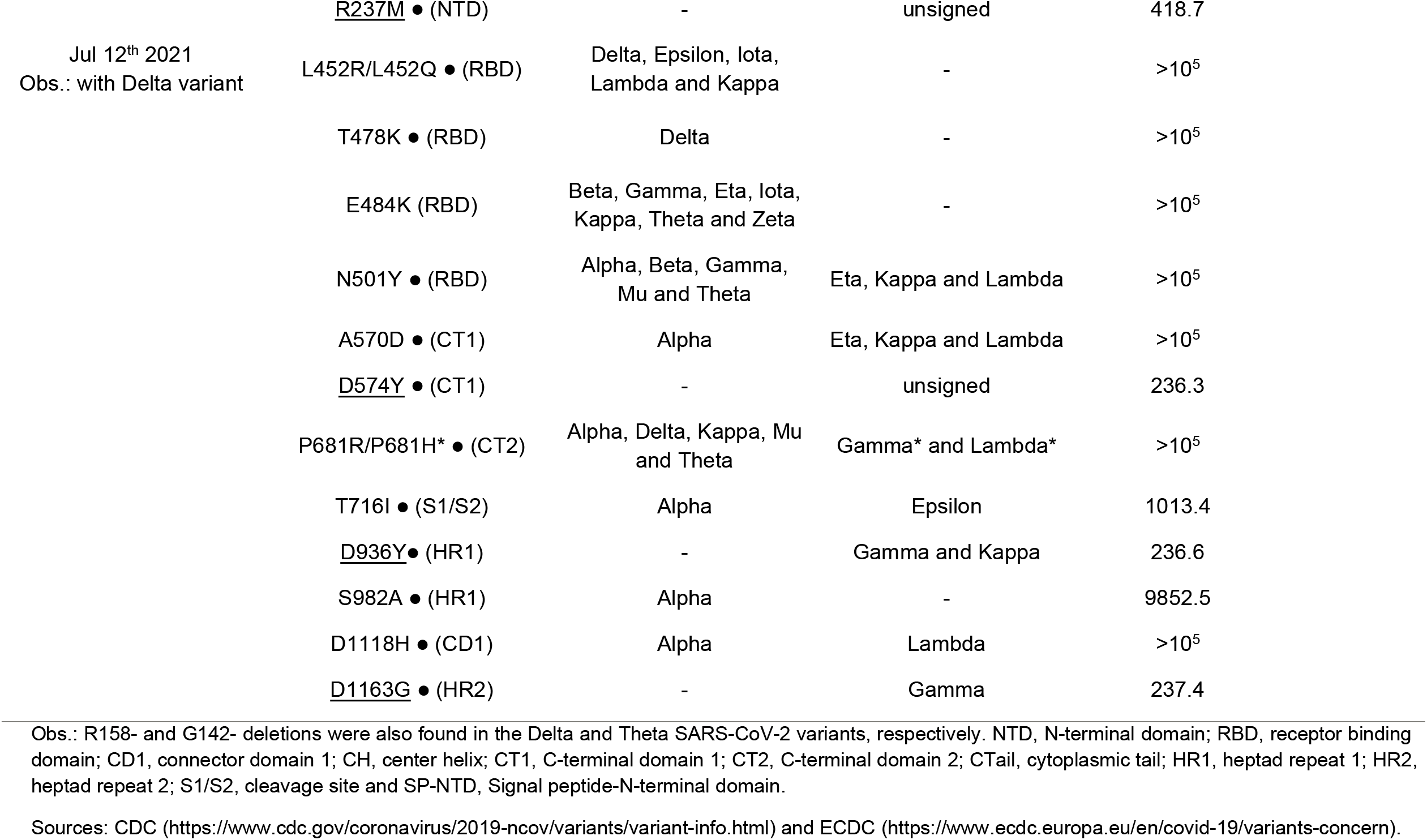
Mutational landscape of SARS-CoV-2 Spike protein VOC and VOI based on the WHO label.

Spike mutations such as E484K, N501Y, L452R, S13I and W152C, seem to be fundamentally important in the process of adaptation of SARS-CoV-2 to human hosts, this by enhancing the affinity to the human ACE2 receptor and mediating immune system evasion (McCallum et al., 2021a; Greaney et al., 2021; Harvey et al., 2021). Our analyzes allowed us to follow SARS-CoV-2 spread dynamics over time in Brazil, showing an increasing number of sites under DEPS, primarily in the Spike-encoding protein. Nine sites are highlighted prior to February 2021, followed by fourteen sites until July 2021 (SARS-CoV-2 Delta variant not included). With the introduction of the Delta variant, both the presence of recombination signals as well as an increase of sites under DEPS were detected (Table 1, Table 2 and Fig. 1), allowing for inferences concerning a SARS-CoV-2 reproductive number increase. An increase in virus circulation augments the chance of viral coinfection, which in turn (and as a pre-requisite for recombination), can heighten the risk of emergence of new variants (Haddad et al., 2020; Ritchie et al., 2021; CDC 2021b).

**Table 2:** Directional evolution landscape on SARS-CoV-2 variants/lineages circulating in Brazil, China, India and the USA.

| **SARS-CoV-2 from** | **Directionally selected residues (proteome)** | **Directionally selected sites (proteome)** | **Most prevalent selection kind in SARS-CoV-2 Spike protein** | **Recombination breakpoints?** |
| --- | --- | --- | --- | --- |
| **Brazil**<br>**(without Delta variant)** | 11 | 89 | Convergent evolution/Repeated Substitutions (10/15 sites) | No |
| **Brazil**<br>**(with Delta variant)** | 16 | 96 | Convergent evolution/Repeated Substitutions (20/22 sites) | Yes |
| **China** | 5 | 19 | Convergent evolution/Repeated Substitution (3/3 sites) | No |
| **India** | 13 | 74 | Convergent evolution/Repeated Substitution (15/15 site) | No |
| **USA** | 20 | 491 | Convergent evolution/Repeated Substitution (12/17 sites) | Yes |

The Delta variant, first identified in late 2020 in India as B.1.617.2 (Kirola 2021), harbors a constellation of non-synonymous mutations in the Spike protein (McCallum et al., 2021b) and has become the leading VOC worldwide. By the end of July 2021, this VOC accounted for 90% of all sequenced samples (Lamarca et al., 2021; PAHO, 2021). Brazil, India and the USA, the countries most severely affected by the pandemic, are now once again threatened by this highly contagious variant. Analysis of the molecular evolution of SARS-CoV-2 taking into account the influence of local demography in these specific scenarios has the potential to generate important insights into the spread and infection dynamics of this pathogen.

Using SARS-CoV-2 sequences from China (the most populated country in the world) as reference, we analyzed all datasets from Brazil, India, and the USA via a large-scale phylogenetic partitions analysis (Prosperi et al., 2011; Matsuda, Suzuki, and Ogata 2020). Increases in SARS-CoV-2 infections were observed to be proportional to locally circulating variants and were not (in the scenarios analyzed), correlated with any particular demography (Fig. 1D); this indirectly reinforces the importance of measures implemented to avoid viral propagation. Analysis of phylogenetic partition clusters along the length of the circa 30 kb CoV genome evidenced several directionally-evolving sites under convergent evolution (Table 2). Thus, a possible association between the rate of infections and the number of residues as well as sites in the Spike-encoding protein under DEPS can be established (Fig. 1D). Interestingly, this supports a hypothesis of convergent evolution due to repeated and multiple site-specific substitutions in distinct SARS-CoV-2 VOC and VOI (see Table 1 and Table 2).

Additionally, we also inferred the possible appearance of SARS-CoV-2 sub-lineages and traced the influence of an environment favoring directional evolution acting on SARS-CoV-2 variants. We showed different patterns among sites in the VOC and VOI, with a particular emphasis on the Kappa VOI currently circulating in the USA. We further demonstrated recombination among SARS-CoV-2 VOC and VOI from India (Beta, Eta and Kappa) and the USA (Beta, Eta and Lambda) (Fig. 2A and Table 2).

**Figure 2.**
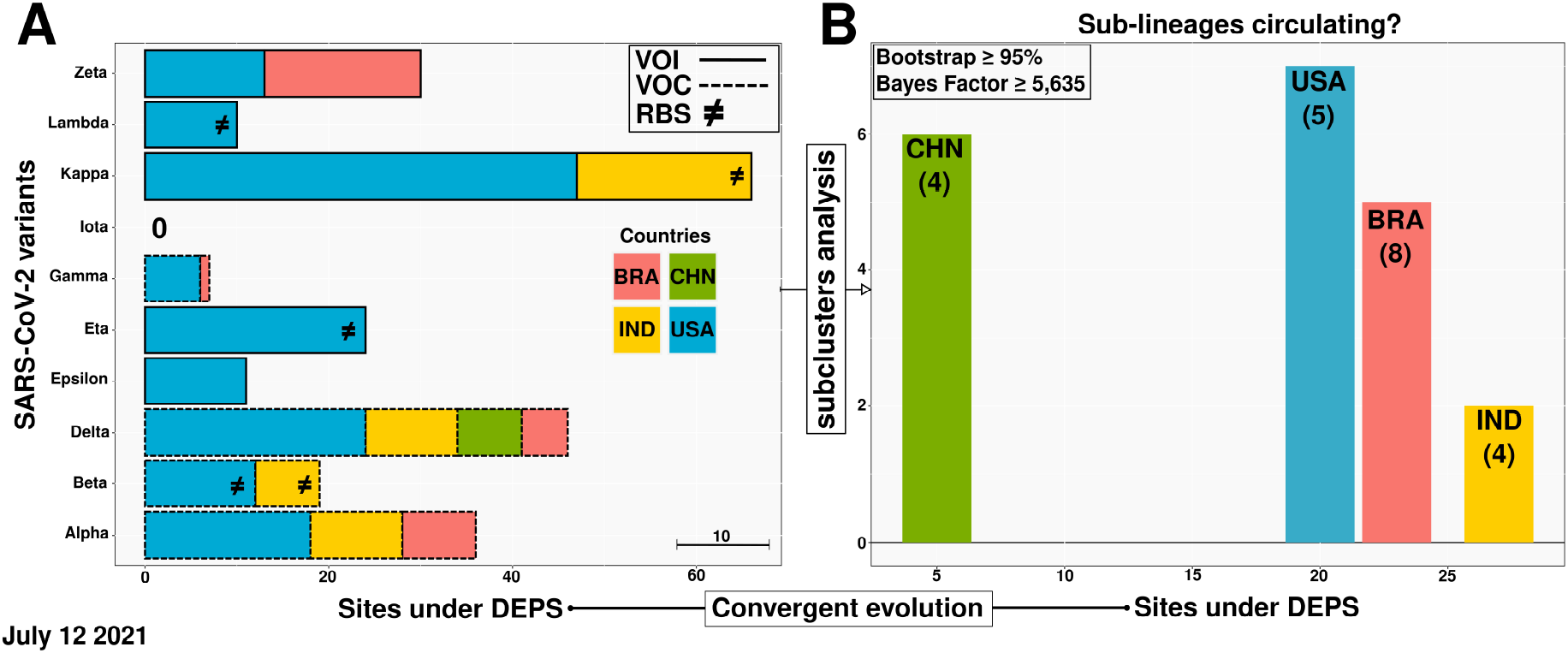
Transmission clustering and sub-clustering analyses on SARS-CoV-2 Variants of Concern (VOC) and Variants of Interest (VOI) sequences. (**A**) Model-based phylogenetic Maximum Likelihood (ML) method to infer transmission clustering and directional evolution in the VOC (dashed line) and VOI (non dashed line), and (**B**) sub-clustering and sub-lineage inferences in strains circulating in Brazil, China, India and the USA. Each color represents a particular country. RBS stands for recombination breakpoints signal (≠) and the scale bar shows the proportion of ten sites under positive selection (A). The numbers in parentheses indicate Spike-encoding protein sites under directional and convergent evolution (B).

As one of the first countries in the world to develop efficient immunizations and implement a vaccination policy (FDA, 2021), the USA vaccinated more than 30% of its population by April 2021. By September 2021, 60% of the booster-immunized population possessed neutralizing antibodies against several viral variants (Ritchie et al., 2021; Pegu et al., 2021). Similar outcomes were observed following widespread vaccination with various SARS-CoV-2 vaccines (different technologies leveraged for vaccine production) in many other regions, including South America and India (Li et al., 2021; Bernal et al., 2021) (Fig. 3). Nonetheless, viral circulation in the face of incomplete immunization has been described as one of the probable causes of the emergence of new variants (Sabino et al., 2021). Accordingly, our own analysis identified SARS-CoV-2 VOC and VOI subclusters (Fig. 2B), thus indicating co-circulation of variants and sub-lineages under convergent evolution. Surprisingly, the same evolutionary pattern was also observed for other endemic and epidemic CoVs studied (see Data availability).

**Figure 3.**
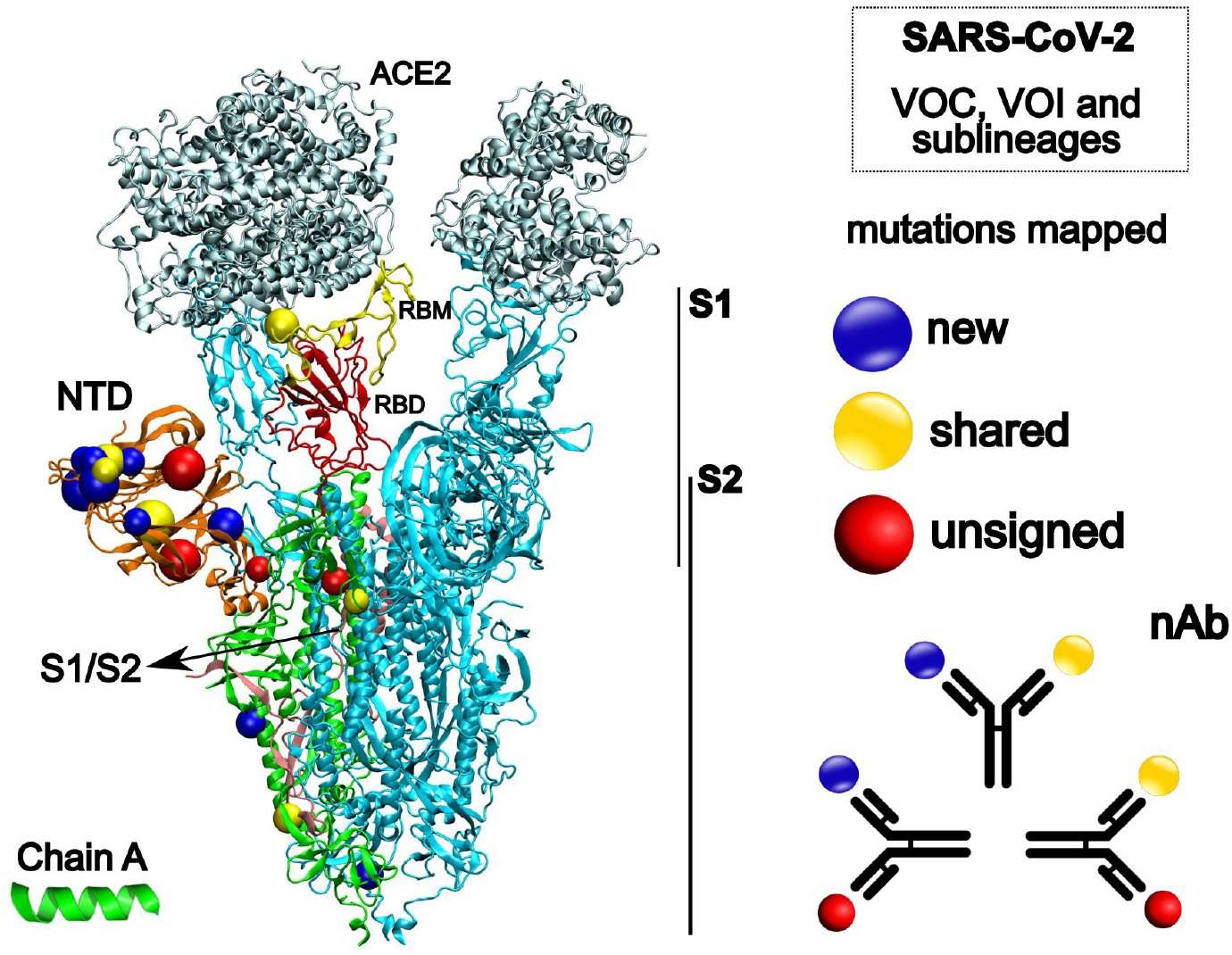
Structural representation of the SARS-CoV-2 Spike glycoprotein (PDB 7A98). On the left side, different colors represent ACE2, angiotensin-converting enzyme 2 (silver); NTD, N-terminal domain (orange); RBM, receptor binding motif (yellow); RBD, receptor binding domain (red) and S1/S2, cleavage site to S2 (pink). The structures in blue represent the chain B and C, respectively. Coloured spheres highlight the mutations mapped in the study. On the right side, our hypotheses about a linear perspective into the prospective vaccine efficacy against different SARS-CoV-2 strains. nAb, neutralizing antibody. Spike protein image was created with the Visual Molecular Dynamics (VMD) v.1.9.3 (Humphrey, Dalke, and Schulten1996).

This study demonstrates the influence of positive directional evolution on SARS-CoV-2 circulating in South America and in those countries most severely affected by the COVID-19 pandemic. Our methodology allowed for the identification of recombination breakpoints and distinct transmission subclusters. We were able to indirectly infer transmission of a viral epidemiological chain and the generation of new variants. We also further identified and classified several convergently emerged shared mutations in different SARS-CoV-2 VOC and VOI. Lastly, we hypothesize that the co-circulation of SARS-CoV-2 variants and their possible sub-lineages takes place within a very close evolutionary environment, which can be translated to a setting of strong convergent evolution, where the viral effective population size have acquired identical site-specific mutations. Our results can help to anticipate a linear perspective with regards to future vaccine efficacy pandemic.

## Data Availability

All data are publicly available at https://github.com/rosadanilo/SARS-CoV-2_DEPS

## Data Availability

Some data on which this paper is based are too large to be retained or publicly archived with available resources. Smaller files which comprise information concerning recombination, convergent evolution, phylogenetic-based clustering analysis (ML trees, transmission clusters/subclusters), as well as the filtered and aligned sequences datasets used to map the shared and unsigned mutations in the SARS-CoV-2 VOC and VOI, are publicly available at https://github.com/rosadanilo/SARS-CoV-2_DEPS.

## Acknowledgments

We gratefully acknowledge the authors and both the originating and submitting laboratories for the sequence data in GISAID EpiCoV and GenBank on which this work is based. The authors also thank the Rede Corona-Ômica/MCTI/FINEP, the National Laboratory for Scientific Computing (LNCC/MCTI, Brazil) for providing HPC resources of the Santos Dumont supercomputer (ID #45691), and Prof. Luiz Mário Ramos Janini for fruitful discussion.

